# Is it the place or the people in the places? Exploration of why young people in deprived coastal communities of England have worse mental health than their peers inland

**DOI:** 10.1101/2025.06.26.25330335

**Authors:** Emily T Murray, Avril Keating, Cara Booker, Claire Cameron, Sam Whewall, Stephen Jivraj

**Affiliations:** Institute of Public Health and Wellbeing, University of Essex, Colchester, CO4 3SQ, UK; Institute of Education, University College London; London, WC1B 7HB, UK; Institute of Social and Economic Research, University of Essex; Colchester; Social Research Institute, University College London, London; Department of Epidemiology and Public Health, University College London; London

## Abstract

Previous research has shown that English adolescents who lived in the most deprived coastal neighbourhoods had worse mental health outcomes up to 11 years later than if they had lived in equivalent inland neighbourhoods. We used the same twelve waves (2009-2022) of Understanding Society, to examine whether environmental characteristics during adolescence, or their socio-demographics, explained this association. All analyses were adjusted for probability of selection into the study via survey weights, clustering of individuals within areas, and attrition over time. During adolescence, coastal youth (n=764) were exposed to worse average levels of sixteen environmental measures and better average levels for five environmental measures, than their peers inland (n=4,157). The concentration of area deprivation was also greater for coastal youth compared with their inland peers. When longitudinal models were fitted between environmental measures and SF-12 mental functioning scores (MCS) during adulthood (age 16+), and adjustments made for individual age, gender, ethnicity, household income and tenure, only local crime and higher education participation were independently associated with MCS scores [Top 20% vs Bottom 20% (95% Confidence interval (CI): −1.20 (−2.38,−0.03) and Middle 20% vs Worse 20%: 1.07 (0.09,2.05)]. However, the amplified effect of area deprivation on MCS scores in coastal, compared to inland, areas was reduced the most by adjustment for individual socio-demographics [interaction term coastal*Top20% deprived area: −5.1 (−8.1,−2.2) to −4.3 (−7.0,−1.6)], then the two environmental measures [further reduced to −3.9 (−6.7,−1.1)]. Interventions to improve the mental health of coastal youth should focus on young people’s socioeconomic circumstances in these areas.

**Funding:** UK Economic and Social Research Council.

## INTRODUCTION

As of 2023, one in five children and young people in England had a probable mental health disorder. This includes 20.3% of 8–16-year-olds, 23.3% of 17–19-year-olds and 21.7% of 20–25-year-olds (1). These percentages have doubled since 2010, with adolescents and young people showing the sharpest increases across multiple national data sets (2). Correspondingly, National Health Service mental health referrals for children and young people have increased by 50% between 2020 and 2023 (3).

However, the occurrence of poor mental health is not distributed equally across England. There is no data on the prevalence of mental health in young people that can provide estimates at the small area level. There are data for adults available from the English small area for mental health index, which clearly show a higher mental health care demand clustered in urban centres and coastal areas (4). The United Kingdom’s (UK’s) Chief Medical Officer (CMO) report from 2021 also highlighted that hospital admissions for self-harm were 35% higher for 10-to 24-year-olds living in coastal, compared to inland, Lower Super Output Areas (LSOAs) (5). When they used GP-level Quality Outcomes Framework (QOF) data from 2014/15 to 2018/19, the coastal excess youth mental health gap was only seen in LSOAs categorised as 7 and 9 of area deprivation (10 being the most deprived). Recent analysis using longitudinal population-level survey data, and self-reported levels of mental functioning and psychological distress, expanded on this finding by showing that the coastal mental health gap only occurred for young adults who had lived in the top 20% most deprived areas of England, not all coastal areas (6). Given that some of the highest rates of suicide in England are in English coastal communities, and that more than two-thirds of mental illness begins by age 25 years (7), it is vital we identify the cause of these elevated levels of poor mental health in coastal youth.

Why young people in the most deprived coastal communities would have worse mental health than their peers in equally deprived inland areas has not been investigated previously. One explanation is that even within the most deprived areas, the environmental factors that cause or mitigate mental health are different in coastal compared to inland areas. The way that area deprivation is constructed is to create one index from multiple indicators that represent material and social disadvantage of residents of small geographical areas. The CMO report uses The English Indices of Deprivation 2019, which uses 39 separate indicators across seven distinct domains (8). One domain is employment deprivation, which is based on the proportion of residents claiming benefits (e.g., jobseeker’s allowance) in August (9). Many coastal communities share a reliance on a seasonal economy, so a data collection at peak tourist season may not accurately reflect coastal residents’ year-round employment conditions. Similar arguments could be made for other indicators. As well, other indicators that have been shown to be related to mental health and potentially more common in coastal youth but are not included in the any of the indexes. For example, social isolation is a growing public health concern that is known to influence mental health (10) and coastal youth are more prone to social isolation from residing in areas with a high retiree populations (5, 11), geographic isolation from other communities (12) and/or public social spaces perceived as only for tourists (13).

Another explanation for why young people in coastal deprived areas would have worse mental health than their peers inland is the high concentration of poverty in these places. Coastal towns were once thriving centres of commerce, but many have experienced stark economic declines in recent decades (14, 15). These economic declines have led to a higher proportion of the population being in poor health, which reduces incentives for business owners to invest in the area, which leads skilled workers to seek employment elsewhere; creating a negative feedback loop of economic and health decline (11). Thus, it is not something about the environments of deprived coastal places that is directly affecting the health of residents (i.e. contextual effects), but more that there are higher concentrations of people in these places who are struggling socioeconomically, and that these individual circumstances are collectively driving poor health outcomes (i.e. compositional effect) (16). There are also indications that levels of deprivation in coastal communities are more severe in coastal than inland areas, with coastal LSOAs making up most of the top 20 rankings of deprivation in the UK (17); but this has not been tested previously. Therefore, the aim of this study is to examine environmental characteristics – here economic, social, educational or built – or individual socio-economic circumstances during adolescence explain the coastal youth mental health gap in the most deprived small areas of England. In order to do so, we test whether: i) average levels of individual and environmental characteristics differ between coastal and inland areas (i.e., exposure)? ii) Are these differences seen within sub-categories of area deprivation (i.e., exposure)? iii) are environmental characteristics associated with mental health in young adulthood (i.e., outcome)? and iv) determine how much of the difference in mental health between deprived coastal youth and deprived inland youth is explained by examined individual and environmental characteristics.

## METHODS

The UK Household Longitudinal Study (UKHLS), also known as *Understanding Society*, is a comprehensive and nationally representative study that tracks the lives of individuals and households across the United Kingdom. The study was launched in 2009-2010 with an initial sample of approximately 40,000 households and has followed up all household members fourteen times, the latest wave in 2021-23 (18).

For this analysis, we focused on data from the youth self-completion questionnaires (ages 10-15) and tracked participants over time to capture their responses to the adult self-completion questionnaires (age 16+). The baseline for each individual was the wave when they completed the youth questionnaire at age 15. In cases where a participant did not complete the questionnaire at age 15, we used the wave closest to that age as their baseline. Response rates among eligible youth varied by wave, with the highest being 82% in wave seven and the lowest 58% in wave 11 (19).

### Measures

#### Coastal community status

Coastal community status was assigned based on the lower-super output area (LSOA) identifier for each youth respondent at their baseline adolescent wave. Each respondent’s usual residence was recorded at every wave, and staff from the Institute for Social and Economic Research (ISER) provided the LSOA identifier for the 2011 Census for all waves (20). In 2011, England contained 32,844 LSOAs, each representing between 400 and 1,200 households, with a typical population ranging from 1,000 to 3,000 individuals (21). These LSOA identifiers were then used to link each youth respondent’s UKHLS data (22) to coastal community status, as defined in the Chief Medical Officer of England’s 2021 report. Briefly, “coastal” LSOAs were those that included or were within 500 meters of built-up areas near the “Mean High Water Mark” (excluding tidal rivers). All other LSOAs in England were classified as “inland.” A more detailed definition can be found in the report (5).

#### Mental health

For this analysis, we used the Mental Component Summary (MCS) score from the 12-item Short-Form Survey (SF-12) as a proxy for mental health. In UKHLS, a measure of psychological distress is also calculated through the General Health Questionnaire (GHQ-12) (23), but MCS scores showed a stronger association with coastal community status in previous analysis (6). In the SF-12, six mental health-related questions were asked about mental well-being in the last four weeks. Answers to these items were converted to a single score by the Ware et al (2002) method (24) calibrated against population norms, by the UKHLS research team. The MCS scores range from 0 (poorest mental health) to 100 (best mental health). Both outcomes were assessed for each respondent at all study waves completed when they were aged 16+ years. The number of possible study waves completed varied by respondent, with a maximum of 12 waves.

#### Covariates

Based on prior research, we included a range of covariates that could be possible alternative explanations for why associations would be seen between coastal community residence and mental health.

Individual level covariates included age at health measurement and six covariates measured in adolescence: gender (self-reported male or female), ethnicity (collapsed into white or non-white), household income (gross monthly income imputed by the ISER team, then adjusted for household size and composition through the OECD-modified equivalence scale (25), consumer price index inflation (26) at that wave and logged) and tenure (collapsed into three categories of homeowner, social renter, or private renter/other). The adolescent measurement corresponds to when coastal community residence was measured for each respondent, ranging from age 10 to 15 years.

Environmental measures were initially considered that could proxy one of the 15 potential pathways Galster theorizes neighbourhoods may affect health of residents (27). Priority was given to measures that had previously been seen in the literature to be associated with mental health (28–32) and to be a particular issue in English coastal communities (5, 11, 12, 15). These mechanisms, or pathways, are outlined in Supplementary Table 1 with the measure identified (if any), a measure of the description, data source and timing of data collection. If multiple time periods were available, the data collection closest to the 2011 census was chosen for consistency.

#### Effect modifier

Area deprivation was previously identified as an effect modifier of the relationship between coastal community status and mental health (6). We used the same measure of area deprivation, the 2011 Townsend Index (z-score summary derived from four census variables: unemployment, non-car ownership, non-home ownership, and overcrowding), at the LSOA-level, when respondents were adolescents. In this analysis, we used the Index that has been divided into quintiles based on the distribution of LSOAs in England (33).

#### Statistical analyses

Initially, comparisons were made across all covariates between coastal community and inland status using analysis of variance for continuous variables and the chi-square statistic for categorical variables. Additionally, the same comparisons were made within each quintile of area deprivation.

To identify the design of the data set, the svyset command in STATA was specified with household number designated as the sampling unit. To account for attrition in the sample over time, we applied the appropriate sample weights from the wave 12 longitudinal weights (34), along with the stratum identifier variable. The weight used, m_indscus_lw, was created by staff at the ISER to match the specific sample: ‘m’ indicates that it refers to the last wave of the analysis, ‘ind’ identifies individual respondents, ‘sc’ denotes the self-completion aspect of the questionnaire, ‘us’ refers to the GPS sample, and ‘lw’ signifies the longitudinal weight applied to the data. The STATA xtset command was then used to specify data to be panel data with more than one wave of data per person.

The main analysis included a series of regression models to examine which environmental variables explain why coastal youth in the most deprived coastal communities had worse mental health on average than their peers in equally deprived communities. All models were fitted using linear regression models with SF-12 MCS scores at the individual and study wave [STATA xi:streg]. To account for clustering of similar individuals within LSOAs, a VCE cluster option was used with LSOA as the cluster variable.

The first series of models aimed to assess which environmental variables were independently associated with mental health in young adulthood. Initially, each environmental measure was fitted separately to assess associations with MCS scores after adjustment for age at MCS score assessment. Following, each separate model was additionally adjusted for adolescent individual covariates. Lastly, independent environmental associations were assessed by fitting all environmental measures jointly with each other and with all individual covariates. The latter model was required as multiple environmental variables were moderately correlated (see Supplementary Table 2).

The second series of models aimed to assess whether environmental measures explained the deprived coastal youth mental health gap. First, MCS scores were regressed on coastal community status, Townsend Index quartiles, a coastal by Townsend interaction term and age at MCS assessment. Second, the individual covariates were added to the model to assess whether associations could be explained by the differences in individual attributes of residents that live in coastal, compared to inland, communities.

Lastly, environmental measures identified as having an independent association with MCS scores in previous analysis were singularly added to the age- and individual covariate-adjusted models.

## RESULTS

Of the 14,746 youths who self-completed a questionnaire at least once, a total of 4,921 youth (18,324 observations, mean=3.7, range 1-11) lived in England at the age of geographic linkage to determine coastal residence, completed at least one adult (age 16+) questionnaire during the follow-up period, and had data on both health outcomes and covariates. Missing data statistics, including excluded and included comparisons, and individual sample characteristics, are available in a previous publication (6). In brief, a higher proportion of coastal adolescent respondents were of White ethnicity and lived in social or private rented homes, compared to those who resided inland.

Table 1 shows the environmental characteristics of respondents by the coastal or inland categorisation. Across the economic, social, educational and built environment domains, coastal adolescent respondents lived in LSOAs with less favourable average levels for sixteen examined factors, compared to their inland peers. This included lower levels of homeownership, NVQ level 3+ qualifications, residents aged 10-19 years, KS4 test scores, participation in higher education, travel time to gambling and tobacco shops, green space within 900m and skilled occupations; as well as higher levels of economic inactivity, urbanicity, mean annual sulphur dioxide and travel times to GPs, further education, large employment centres, job centres, and leisure. For seven environmental variables, coastal respondents lived in LSOAs with more favourable average levels than respondents in inland LSOAs with regards deprivation, crime deprivation, proportion households overcrowded, walking distances to food stores and levels of air pollution (annual nitrogen dioxide and particulate matter 10).

**Table 1.**
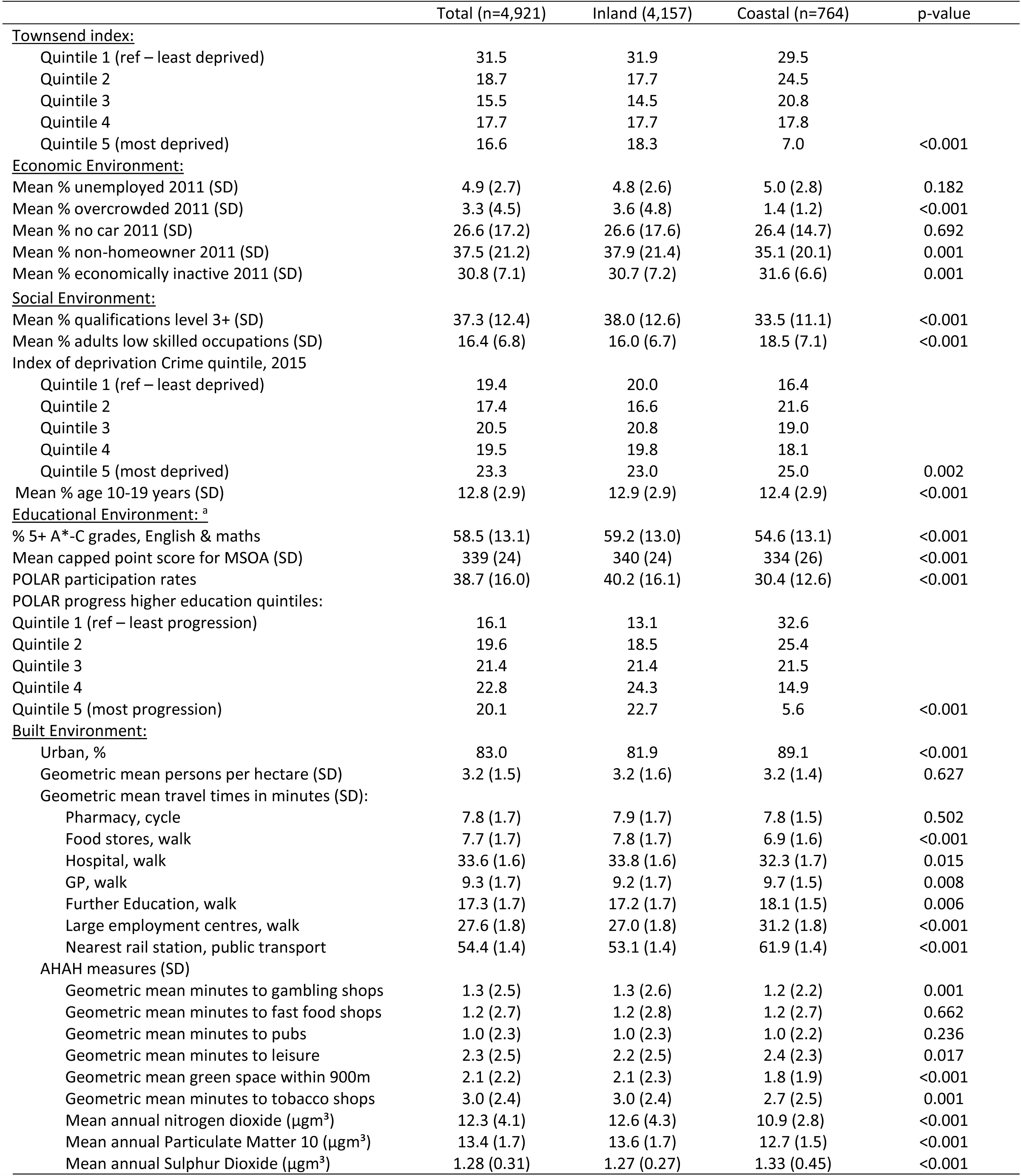
Distribution of environment variables for analysis sample: all, Inland community, Coastal community, UKHLS youth sample, 2009-2021 (n=4,961).

Even when the sample was split further by area deprivation categories (see Table 2), there were consistent differences in environmental variables between coastal and inland LSOAs. For the UKHLS young adults who lived within the most deprived quartile of English LSOAs during adolescence, those in coastal areas had less favourable levels of most social, economic, and educational environmental measures than inland areas in the same quintile categorisation of deprivation. This included a lower proportion of adults obtaining Level 3 qualifications, more adults in lower skilled occupations, higher average crime deprivation, higher unemployment, lower average GCSE attainment scores and lower prevalence of progress to higher education. The largest differences were in the educational measures with, for example, 21% of 18–19-year-olds in the most deprived coastal MSOAs participated in higher education, while 35.8% of their inland peers in the most deprived category participated. For built environment measures, within deprivation-category differences between coastal and inland LSOAs were significant, but small (average ∼1 minute difference). Similar patterns were seen when all English LSOAs were examined, not just those where UKHLS respondents resided (See supplementary Table 3). This indicates there is no geographic selection bias in the sampling strategy of the UKHLS by whether an LSOA is coastal or not.

**Table 2:**
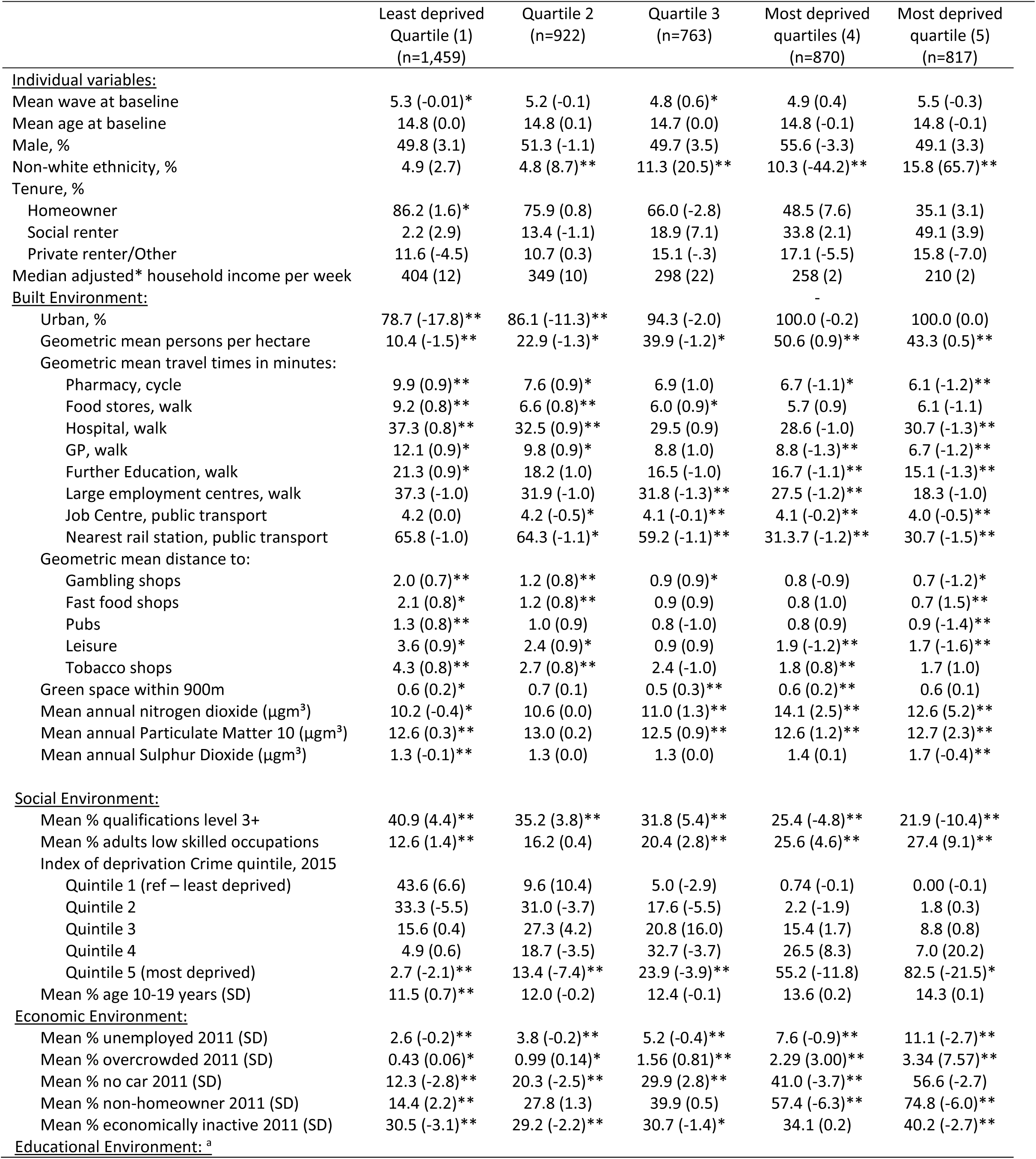
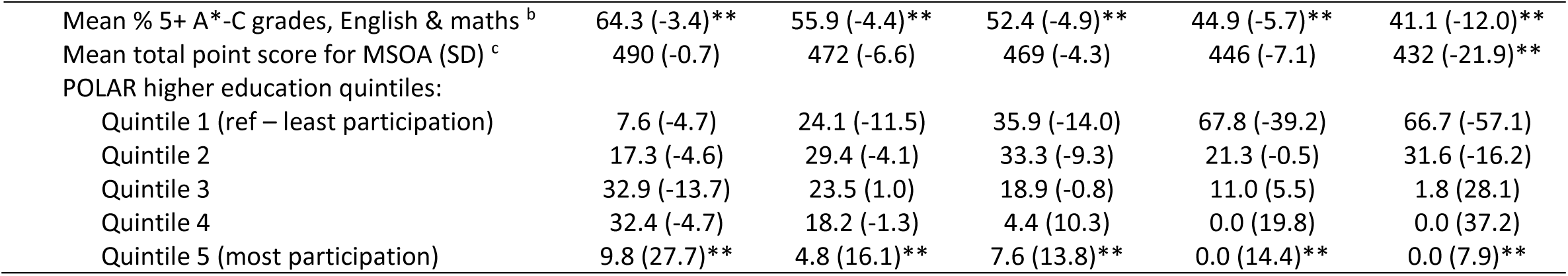
Average individual and environmental variables for UKHLS youth sample members who lived in Coastal lower-super output areas (difference Inland - Coastal areas) by Area deprivation quartiles, 2009-2021 (n=4,961).

However, only a small selection of environmental variables were associated with SF-12 mental functioning component (MCS) scores in age-adjusted models (see Table 3). Most economic measures were associated with higher (better) MCS scores, but most were subsequently explained by adjustment for individual socio-demographic variables, except the proportion of households with no car [0.10 (95% CI: 0.02, 0.18]. For the social and educational domains, only the higher the proportion 10–19-year-olds and quintile three of higher education progression (vs quintile 1), were associated with higher MCS scores in both age- and socio-demographic-adjusted models. However, for areas in the highest crime quartile, compared to lowest, associations were only seen after adjustment for socio-demographic variables [-1.33 (95% CI: −2.26, −0.41)]. A similar pattern was seen for the built environment domain, where longer travel times to employment and job centres was associated with lower (worse) MCS scores, and higher levels of nitrogen dioxide and particulate matter 10 were associated with higher MCS scores, but these were explained by adjustment for individual socio-demographics. While associations between higher travel times to pubs and leisure were associated with lower (worse) MCS scores, these were only apparent after adjustment for individual socio-demographics. The only environmental variables to show independent associations with MCS scores in the full model (table 3, model 3) were the highest crime quartile [-1.20 (95% CI: −2.38, −0.03) and the middle quartile of participation in higher education [1.07 (0.09, 2.05)].

**Table 3:** Estimated mean SF-12 mental functioning component (MCS) score by 1-unit change in environmental variables, UKHLS youth sample members, 2009-2021 (n=4,961).

|  |  |  |  |
| --- | --- | --- | --- |
| Mean % unemployed 2011 (SD) | **0.14 (0.03, 0.25)** | -0.01 (-0.13, 0.11) | - |
| Mean % overcrowded 2011 (SD) | **0.19 (0.12, 0.25)** | 0.03 (-0.01, 0.07) | - |
| Mean % no car 2011 (SD) | **0.02 (0.00, 0.04)** | **0.10 (0.02, 0.18)** | 0.02 (-0.01, 0.05) |
| Mean % non-homeowner 2011 (SD) | 0.01 (-0.002, 0.02) | -0.01 (-0.03, 0.01) | - |
| Mean % economically inactive 2011 (SD) | **0.08 (0.04, 0.12)** | -0.01 (-0.02, 0.01) | - |
| <u>Social Environment:</u> |  |  |  |
| Mean % qualifications level 3+ (SD) | -0.02 (-0.04, 0.005) | -0.01 (-0.03, 0.02) | - |
| Mean % adults low skilled occupations (SD) | 0.01 (-0.04, 0.05) | -0.01 (-0.05, 0.04) | - |
| Index of deprivation Crime quintile, 2015 |  |  |  |
| Quintile 1 (ref – least deprived) | - | - | - |
| Quintile 2 | -0.72 (-1.63, 0.19) | -0.83 (-1.70, 0.04) | -0.70 (-1.58, 0.18) |
| Quintile 3 | 0.03 (-0.85, 0.91) | -0.31 (-1.18, 0.54) | -0.06 (-1.00, 0.88) |
| Quintile 4 | 0.57 (-0.29, 1.42) | -0.22 (-1.09, 0.65) | -0.03 (-1.03, 0.97) |
| Quintile 5 (most deprived) | -0.16 (-1.04, 0.72) | **-1.33 (-2.26, -0.41)** | **-1.20 (-2.38, -0.03)** |
| Mean % age 10-19 years (SD) | **0.17 (0.08, 0.26)** | **0.10 (0.01, 0.19)** | 0.09 (-0.004, 0.19) |
| <u>Educational Environment:</u><sup>a</sup> |  |  |  |
| % 5+ A*-C grades, English & maths | 0.0 (-0.02, 0.02) | 0.01 (-0.01, 0.04) | - |
| Mean capped point score for MSOA (SD) | 0.0 (-0.01, 0.01) | 0.00 (0.00, 0.01) | - |
| POLAR participation rates | 0.01 (-0.01, 0.03) | 0.01 (-0.01, 0.02) | - |
| POLAR progress higher education quintiles: |  |  |  |
| Quintile 1 (ref – least progression) | - | - | - |
| Quintile 2 | 0.38 (-0.60, 1.36) | 0.21 (-0.76, 1.17) | 0.20 (-0.79, 1.20) |
| Quintile 3 | **1.51 (0.56, 2.46)** | **1.07 (0.12, 2.01)** | **1.07 (0.09, 2.05)** |
| Quintile 4 | 0.85 (-0.07, 1.77) | 0.57 (-0.36, 1.50) | 0.65 (-0.34, 1.64) |
| Quintile 5 (most progression) | 0.85 (-0.09, 1.79) | 0.62 (-0.33, 1.57) | 0.75 (-0.30, 1.80) |
| <u>Built Environment:</u> |  |  |  |
| Urban, % | 0.05 (-0.71, 0.82) | -0.47 (-1.21, 0.28) | - |
| Geometric mean persons per hectare (SD) | 1.12 (0.93, 1.34) | 0.86 (0.71, 1.04) | - |
| Geometric mean travel times in minutes (SD): |  |  |  |
| Pharmacy, cycle | 0.59 (0.34, 1.03) | 1.04 (0.59, 1.85) | - |
| Food stores, walk | 1.26 (0.74, 2.14) | 1.68 (1.00, 2.84) | - |
| Hospital, walk | 0.62 (0.36, 1.09) | 1.12 (0.63, 2.00) | - |
| GP, walk | 0.63 (0.37, 1.08) | 1.14 (0.65, 2.00) | - |
| Further Education, walk | 0.98 (0.57, 1.70) | **0.77 (1.00, 3.12)** | - |
| Large employment centres, walk | **0.58 (0.36, 0.93)** | 0.98 (0.60, 1.61) | - |
| Nearest job centre, public transport | **0.27 (0.12, 0.59)** | 0.97 (0.40, 2.36) | - |
| AHAH measures (SD) |  |  |  |
| Geometric mean minutes to gambling shops | 0.80 (0.59, 1.08) | 1.17 (0.86, 1.60) | - |
| Geometric mean minutes to fast food shops | 0.82 (0.62, 1.09) | 1.21 (0.90, 1.63) | - |
| Geometric mean minutes to pubs | 1.09 (0.79, 1.52) | **1.46 (1.06, 2.02)** | 1.36 (0.88, 2.09) |
| Geometric mean minutes to leisure | 0.93 (0.68, 1.27) | **1.41 (1.01, 1.95)** | 1.19 (0.77, 1.86) |
| Mean green space within 900m | -0.07 (-0.37, 0.24) | -0.08 (-0.39, 0.22) | - |
| Geometric mean minutes to tobacco shops | -0.01 (-0.33, 0.32) | 1.33 (0.96, 1.85) | - |
| Mean annual nitrogen dioxide (µgm<sup>3</sup>) | **0.16 (0.09, 0.23)** | 0.05 (-0.03, 0.13) | - |
| Mean annual Particulate Matter 10 (µgm<sup>3</sup>) | **0.18 (0.01, 0.34)** | 0.00 (-0.18, 0.18) | - |
| Mean annual Sulphur Dioxide (µgm<sup>3</sup>) | 0.90 (-0.04, 1.85) | 0.67 (-0.25, 1.58) | - |

Table 4 shows associations between coastal residence, area deprivation (measured by Townsend index) and coastal*Townsend quintiles before and after adjustment for the individual socio-demographic and environmental variables. In age-adjusted models, respondents who had lived in one of the most deprived coastal communities in adolescence, their mean MCS scores in young adulthood were −5.1 points lower (95% CI: −8.1, −2.2), compared to the least deprived inland communities (table 4, model 1). The separate addition of socio-demographic covariates and higher education progression reduced this amplification effect to −4.3 (−7.0, −1.4) and −4.7 (−7.7, −1.7), respectively, while adjustment for local crime levels (and an interaction between coastal and crime) increased the association to −6.1 (−9.7, −2.4). Inclusion of both environmental variables improved model fit slightly (R^2^ 0.0182 to 0.0198 and 0.0196). The final model inclusive of all individual and environmental variables, showed that adolescents residing in the top 20% most deprived coastal communities had MCS scores on average −3.9 points (95% CI: −6.7, −1.1) lower than peers in the least 20% deprived inland communities.

**Table 4:**
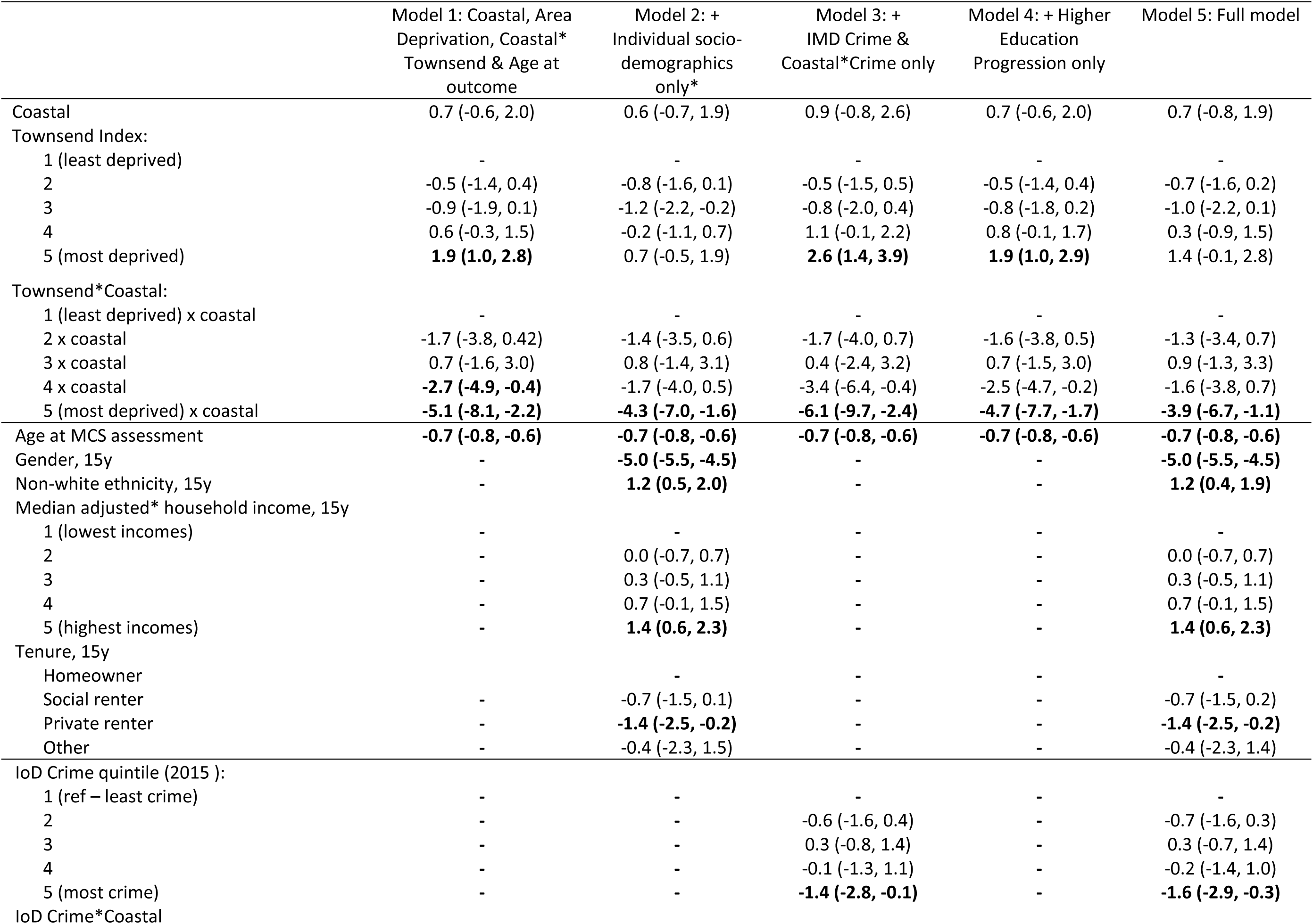
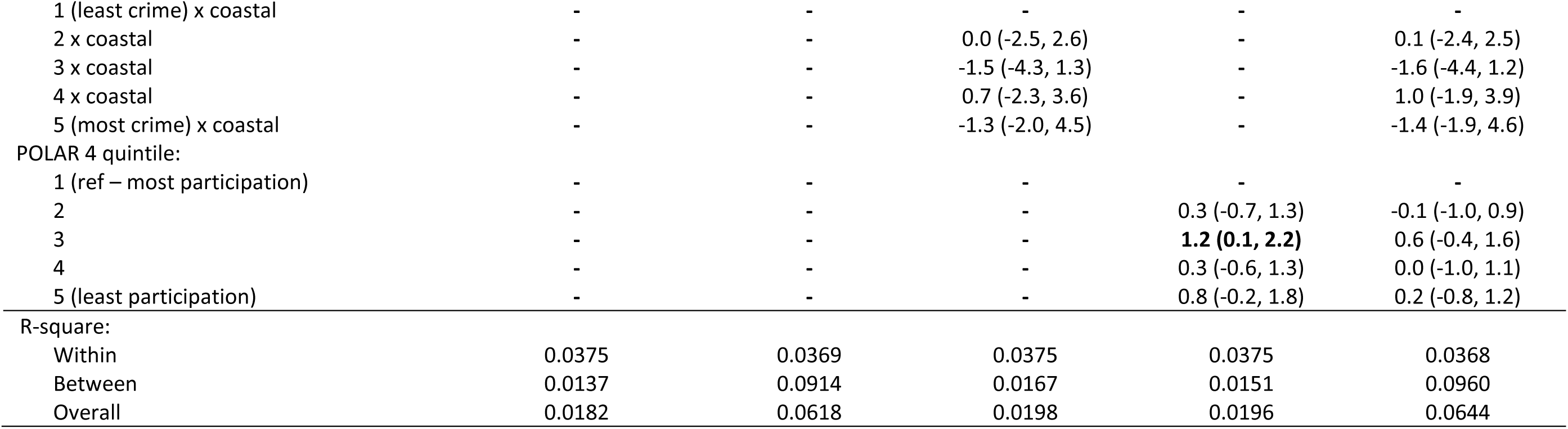
Adjusted associations of SF-12 mental functioning score (95% CI) in young adulthood across 11 waves of follow-up, by environmental factors in adolescence, UKHLS youth sample, 2009-2021 (n= n=4,961, observations=18,324): Townsend index as quintiles.

## DISCUSSION

Using a nationally representative sample of adolescents who lived in England from 2009 to 2020, and up to eleven years of follow-up, we have shown that young adults who had lived on the coasts of England during adolescence were living in neighbourhoods that had worse levels of multiple economic, social, educational and built environmental factors known to be associated with poor mental health, than their inland peers. This remained the case when comparisons were made between coastal and inlands adolescents living in the most deprived fifth of neighbourhoods. When these young people were followed up into young adulthood, their mental functioning was independently related to both the level of crime and participation in higher education there had been in their adolescent neighbourhoods. However, in further analysis, these two environmental measures did not substantially explain differences in mental functioning between deprived coastal and inland youth: our main focus of enquiry. Adjustment for respondent’s demographic and socioeconomic circumstances reduced associations the most (∼16%), but the mental health gap remained after adjustment for all included individual and environmental factors.

The finding that youth respondents who lived in coastal neighbourhoods were exposed to more adverse levels of a host of environmental variables, than their inland peers, is consistent with numerous reports on English coastal communities (5, 11, 12, 15). There is a common story, which our data supports, that many coastal communities share characteristics of struggling labour markets, lower skilled populations, lower educational attainment and lower homeownership (14). However, in terms of distances to local health-harming and health-promoting infrastructure, the built environments where coastal adolescents lived ∼10 years ago were not hugely different to where their peers lived inland. As well, there was a positive story that coastal youth were exposed to appreciably lower levels of outdoor air pollution, namely No2 and PM10; which has been linked to increased risk of depressive symptoms and incidence of suicide (35, 36). In addition to the above reports, we concernedly show that even within the top 20% most deprived neighbourhoods in England, coastal youth were living in areas with worse economic, social and educational environments than their equivalent peers inland, showing that deprived coastal areas should be prioritised for national and local government regeneration initiatives.

Our third finding that local area crime and participation in higher education (POLAR) were independently associated with mental health is half supported by previous studies. There is a large literature showing robust evidence that higher levels of crime are related to higher levels of depression and psychological distress, even after adjustment for individual socio-demographics (37). This is seen to occur through direct impact on mental health of being a victim or witness of a crime, or indirectly though feeling that particular places are unsafe creating a biological stress response and/or change behaviour(s) that effect health (e.g., less physical activity or social engagement in unsafe areas) (38). In our study, the main effect of local area crime was robust to adjustment for coastal residence, area deprivation and all individual socio-demographics. There was also an indication of an amplification of the relationship between local crime rates and mental functioning for coastal, compared to inland, residents; but this was explained by socio-demographics of youth participants.

For the POLAR measure, as far as we are aware this is the first study to show that young people in the middle quintile of the POLAR measure had slightly better mental functioning scores than those in areas where the highest proportion of young people participate in higher education. The hypothesized mechanism for why neighbourhood higher education progression was linked to individual mental health was one of social contagion. That seeing other young people in your neighbourhood progressing to higher education would build confidence in your own ability to progress and do well academically (39, 40). However, as this association did not increase with additional levels of participation, and most of this association was reduced by adjustment for individual socio-demographics, it is likely that this finding is explained by the individual attributes of study youth who resided in these neighbourhoods.

Our results showing that only two built environmental measures - distance to pubs and leisure centres – were related to mental functioning in young adulthood is somewhat consistent with a similar aged New Zealand study (30). They showed in a cross-sectional design that young people aged 10-24 years who lived in ‘health-constraining’ environments had higher odds of any diagnosed mental health condition. Why we did not see similar findings across our other ‘health-constraining’ environmental measures could be due to reverse temporality issues in the NZ study. We used twelve waves of longitudinal data in our analysis, so can be sure of temporal order of relationships. Our associations were also entirely explained by adjustment for individual socio-demographics, suggesting that at least in England, built environment associations with mental health are due to clustering of similar kinds of people residing within the same neighbourhoods, rather than infrastructure. Surprisingly, we did not see an association between availability of green space and youth mental health, which has been seen in some previous studies (41–43). However, the overall literature is mixed with one systematic review suggesting positive findings were due to selection bias and residual confounding (44).

It was therefore not too surprising that the coastal amplification of the area deprivation and mental functioning relationship was hardly explained by the inclusion of local higher education participation or local crime rates. Model fit was slightly improved by inclusion of the two environmental measures. However, our results indicate that a large part of the explanation is the economic and social challenges that people face who live in these communities. Household incomes and private renting are key factors. Many current deprived coastal towns have experienced stark economic declines in recent decades due to the decline of seaside resorts, fishing, and shipbuilding industries (45).

This has created a negative feedback loop of highly skilled and healthy individuals leaving these areas for better employment opportunities (11, 13) leaving behind the sicker and more socio-demographically challenged (46). There is however no research on how much internal migration contributes to the relationship between coastal residence and health outcomes. It is also worth noting that this pattern where individual measures explain more of a health inequality than area-level measures are common in the area-effects literature (47). It does not mean that where people live is not important for their health. The collective interactions, behaviours and decision-making of people who live in neighbourhoods helps to create the ‘context’ for their neighbours and future residents (48). It should also be pointed out that even after we adjusted for all potentially explanatory factors in our study, there was still a sizable difference in mental functioning between deprived coastal and inland young adults. Further work is needed to identify other potential explanations.

One such reason could be due to one of the limitations of our study, in that we only assessed environmental measures at one time point, during adolescence. Previous research has shown that the residential environment has a stronger association with health outcomes at the time of measurement (49). However, since two-thirds of mental illness begin before the age of 25 years (7), we thought it important to examine environmental exposures around the likely time of mental illness development. Research has also shown that people with mental health problems are more likely to move to deprived neighbourhoods (50). Therefore, by focusing on the relationship of adolescent environments with young adult mental health, we reduce issues from reverse temporality and health selection bias. The reason that none of the environmental measures substantially explained differences in mental functioning between deprived coastal and inland youth could be that we did not include the right environmental measure, but it could also be due to data availability issues of most environmental measures not being measured at exactly the time of adolescent residence or exposures occurred elsewhere, such as school environment (i.e., measurement error). As in any longitudinal study, attrition occurred over follow-up, particularly the transition from the youth to adult surveys (6). However, attrition bias is most likely to create underestimates of neighbourhood effects on health over time, given that cohort members residing in the most deprived neighbourhoods, with the worse mental functioning, would be the most likely to leave this study (51).

The major strengths of our study were the ability draw upon a nationally representative, longitudinal, sample of youth that were linked to granular administrative data. This allowed us to be confident that results are generalisable to all English youth during the study period, that we have a comprehensive picture of the environments that these youth resided during their adolescence and that results are unlikely to be the result of reverse causality. The latter point buoyed by previous results showing that only 2% of the sample moved between coastal and inland communities between adolescence and young adulthood (6). Couple this with the comprehensive socio-economic data collected on UKHLS youth participants, and the rest of their household, we are confident results are robust.

In conclusion, findings from this study suggest that strategies to improve mental health of English youth needs to pay particular attention to the socio-demographics of young people, and their families, in deprived coastal communities. Suicide rates and deaths of despair, which include suicide but also those attributable to alcohol or drugs, are significantly higher in coastal compared to inland local authorities (52). Rates of deaths of despair are highest at mid-life (52), with the suicide rate in the UK the highest it has been in over 25 years (53). But given the increasing prevalence of youth mental disorders in England (1), and that more than two-thirds of mental illness begins by age 25 years (7), resources should be targeted in today’s youth to prevent a tsunami of future mental ill health and suicides.

## Funder

United Kingdom Economic and Social Research Council.

## Data Availability

All data produced in the present study are available upon reasonable request to the UK Data Service.

https://ukdataservice.ac.uk/

