## Supplemental files for "Is it the place or the people in the places? Exploration of why young people in deprived coastal communities of England have worse mental health than their peers inland"

**Supplementary Table 1.** Description of potential environmental mechanisms with the proxy measurement identified (if one), a description of the measure (including source and time period) and relevance to coastal communities and mental health.

| Domain | Mechanism | Measure(s) identified | Description of measure | Source (collection period) | Coastal community issue? | Linked to mental health? |
|---|---|---|---|---|---|---|
| **Social-Interactive Mechanisms** | Social Contagion | Educational attainment and progression | • Average Key Stage 4 (GCSE or equivalent) total scores for all eligible pupils in a middle layer super output area (MSOA).<br>• POLAR4 assigns a quintile to each middle-super output area based on how many young people from that MSOA started a higher education course. | • National Pupil Database (2011)<br>• Office for Students. The Cohort started a course between 2009-10 and 2013-14. | Some | Y |
| | Collective Socialization | Neighbours with qualifications, skilled occupations and economically active | • Proportion residents with Level 3 qualifications (requirement for entry to higher education or skilled employment).<br>• Proportion occupied persons in the LSOA with NS-SEC classes 1 (Higher managerial, administrative and professional occupations), 2 (Intermediate occupations) or 3 (Small employers and own account workers), out of five classes.<br>• Proportion of usual residents aged 16 to 74 in the LSOA who were economically active on census day. | Derived from UK Census (2011) | Some | Y |
| | Social Networks | Social isolation | Proportion of population in an LSOA that is aged 10-19 years. | UK Census (2011) | Y | Y |

| | | | | | | |
|---|---|---|---|---|---|---|
| | Social cohesion and control | None | - | - | N | Y |
| | Competition | Housing availability and affordability | Median price paid for lower layer super output areas | Office for National Statistics (2009-2021) | Some | Y |
| | Relative Deprivation | Townsend Index | z-score summary derived from four census variables: proportion unemployed, proportion non-car owners, proportion non-homeowners, and proportion overcrowded households in each LSOA. | Derived from UK Census (2011) | Some | Y |
| | Parental Mediation | None | - | - | Y | Y |
| **Environmental Mechanisms** | Exposure to Violence | Local area crime | Rate of recorded crime in an area for four major crime types representing the risk of personal and material victimisation at a small area level. | Index of Multiple Deprivation Crime score (2015) | Some | Y |
| | Physical Surroundings | Urbanicity and population density | • Rural/urban classification<br>• Persons per hectare | • ONS postcode directory (2011)<br>• Derived from Census (2011) | Some | Y |
| | Toxic Exposure | Air pollution | • Annual mean nitrogen dioxide (µgm3)<br>• Annual mean particulate Matter 10 (µgm3)<br>• Annual mean Sulphur Dioxide (µgm3) | Access to Healthy Assets & Hazards (AHAH) dataset, version 2 (2015) | Some | Y |
| **Geographical Mechanisms** | Spatial Mismatch | Distance to employment centres | Average travel time in minutes to employment centre by walking | Department for Transport (2015) | Y | Y |
| | Public Services | Distance to hospitals, doctors’ offices and job centres | • Average travel time in minutes to nearest hospital by walking<br>• Average travel time in minutes to nearest GP by walking | Department for Transport - journey time statistics (2015) | Y | Y |

| | | | | | | |
|---|---|---|---|---|---|---|
| | | | • Average travel time in minutes to employment centre by walking | | | |
| **Institutional Mechanisms** | Stigmatization | None | - | - | Y | Y |
| | Local Institutional Resources | Distance to further education and green space | Total green space areas available to each postcode in a range of a 900-meter buffer (passive) before creating LSOA level averages. | Access to Healthy Assets & Hazards (AHAH) dataset, version 2 (2015) | Y | Y |
| | Local Market Actors | Distance to pharmacies, food stores, gambling shops, fast food, pubs, leisure and tobacco shops | • Mean travel times in minutes by walking (exception pharmacies is cycling). | Department for Transport - journey time statistics (2015) for pharmacies and food stores. Other measures from Access to Healthy Assets & Hazards (AHAH) dataset, version 2 (2015) | Some | Y |

**Supplementary Table 2.** Correlations of environmental measurements during adolescence.

| | Inctv | Qul3 | Skill | 10-19 | Urb an | PPH | Phrm | Food | Hsp | GP | FE | EmC | JbC | Gmbl | FF | Pubs | Leis | Green | Tbc | No | PM | So | KS4 |
|---|---|---|---|---|---|---|---|---|---|---|---|---|---|---|---|---|---|---|---|---|---|---|---|
| Inactve | 1.0 | | | | | | | | | | | | | | | | | | | | | | |
| Qual3 | -0.5 | 1.0 | | | | | | | | | | | | | | | | | | | | | |
| Skill | -0.4 | 0.9 | 1.0 | | | | | | | | | | | | | | | | | | | | |
| 10-19y | 0.4 | -0.3 | -0.2 | 1.0 | | | | | | | | | | | | | | | | | | | |
| Urban | -0.1 | 0.2 | 0.2 | -0.1 | 1.0 | | | | | | | | | | | | | | | | | | |
| PPH | 0.2 | -0.2 | -0.2 | 0.2 | -0.7 | 1.0 | | | | | | | | | | | | | | | | | |
| Pharm | -0.2 | 0.2 | 0.2 | -0.1 | 0.5 | -0.7 | 1.0 | | | | | | | | | | | | | | | | |
| Food | -0.1 | 0.2 | 0.2 | 0.0 | 0.5 | -0.6 | 0.6 | 1.0 | | | | | | | | | | | | | | | |
| Hosp | -0.2 | 0.1 | 0.1 | -0.1 | 0.4 | -0.5 | 0.5 | 0.4 | 1.0 | | | | | | | | | | | | | | |
| GP | 0.2 | 0.1 | 0.04 | -0.1 | 0.4 | -0.6 | 0.7 | 0.6 | 0.4 | 1.0 | | | | | | | | | | | | | |
| FE | -0.1 | 0.03 | 0.05 | -0.1 | 0.5 | -0.6 | 0.5 | 0.5 | 0.4 | 0.5 | 1.0 | | | | | | | | | | | | |
| EmpC | -0.1 | 0.01 | -0.0 | -0.1 | 0.5 | -0.5 | 0.5 | 0.4 | 0.5 | 0.5 | 0.5 | 1.0 | | | | | | | | | | | |
| JbC | -0.3 | 0.03 | 0.2 | -0.1 | 0.5 | -0.6 | 0.6 | 0.4 | 0.5 | 0.6 | 0.5 | 0.6 | 1.0 | | | | | | | | | | |
| Gmbl | -0.2 | 0.2 | 0.1 | -0.1 | 0.7 | -0.8 | 0.7 | 0.6 | 0.5 | 0.6 | 0.6 | 0.5 | 0.6 | 1.0 | | | | | | | | | |
| FF | -0.2 | 0.1 | 0.0 | -0.1 | 0.6 | -0.7 | 0.7 | 0.5 | 0.5 | 0.6 | 0.6 | 0.5 | 0.6 | 0.8 | 1.0 | | | | | | | | |
| Pubs | -0.1 | -0.1 | 0.1 | 0.04 | 0.5 | -0.7 | 0.6 | 0.5 | 0.4 | 0.5 | 0.5 | 0.5 | 0.6 | 0.7 | 0.8 | 1.0 | | | | | | | |
| Leis | -0.1 | 0.00 | 0.1 | -0.0 | 0.6 | -0.6 | 0.6 | 0.5 | 0.5 | 0.5 | 0.5 | 0.5 | 0.6 | 0.7 | 0.7 | 0.6 | 1.0 | | | | | | |
| Grn | -0.1 | 0.09 | 0.2 | -0.0 | 0.0 | 0.0 | 0.0 | 0.1 | 0.0 | 0.0 | 0.0 | 0.0 | 0.6 | 0.0 | 0.1 | 0.0 | 0.0 | 1.0 | | | | | |
| Tbac | -0.2 | 0.1 | 0.2 | -0.1 | 0.6 | -0.6 | 0.5 | 0.4 | 0.4 | 0.4 | 0.4 | 0.5 | 0.0 | 0.6 | 0.7 | 0.6 | 0.6 | 0.1 | 1.0 | | | | |
| No2 | 0.2 | 0.00 | 0.1 | 0.1 | -0.5 | 0.6 | -0.5 | -0.4 | -0.4 | -0.5 | -0.5 | -0.5 | 0.5 | -0.6 | -0.6 | -0.5 | -0.6 | 0.0 | -0.4 | 1.0 | | | |
| PM10 | -0.03 | 0.08 | 0.2 | 0.0 | -0.2 | 0.4 | -0.3 | -0.2 | -0.2 | -0.3 | -0.4 | -0.2 | -0.9 | -0.3 | -0.4 | -0.3 | -0.3 | 0.0 | -0.2 | 0.7 | 1.0 | | |
| So2 | 0.2 | -0.3 | -0.3 | 0.1 | -0.2 | 0.2 | -0.2 | -0.2 | -0.1 | -0.1 | -0.1 | -0.2 | -0.4 | -0.2 | -0.1 | -0.1 | -0.1 | 0.1 | -0.2 | 0.2 | 0.0 | 1.0 | |
| KS4 | -0.1 | 0.2 | 0.3 | -0.1 | 0.0 | -0.1 | 0.1 | 0.1 | 0.0 | 0.1 | 0.0 | 0.0 | 0.0 | 0.1 | 0.1 | 0.1 | 0.1 | 0.0 | 0.1 | 0.0 | -0.1 | 0.1 | 1.0 |
| PLR | -0.2 | 0.7 | 0.7 | -0.2 | 0.2 | -0.1 | 0.1 | 0.2 | 0.0 | 0.0 | 0.0 | 0.0 | -0.1 | 0.1 | 0.0 | 0.0 | 0.0 | 0.1 | 0.1 | 0.1 | 0.1 | -0.3 | 0.3 |

**Supplementary Table 3: Average individual and environmental variables for English Coastal lower-super output areas (difference Inland - Coastal areas) by Area deprivation quartiles, 2011 (n=32,844)**

| | Least deprived Quartile (1) (n=11,289) | Quartile 2 (n=7,064) | Quartile 3 (n=5,639) | Most deprived quartiles (4) (n=5,426) | Most deprived quartile (5) (n=3,426) |
|---|---|---|---|---|---|
| Built Environment: | | | | - | |
| Urban, % | 82.2 (-19.6)** | 87.3 (-8.1)** | 96.3 (-3.2)** | 98.5 (0.3) | 100.0 (-0.03) |
| Geometric mean persons per hectare | 13.6 (-6.5)** | 25.5 (-7.8)** | 41.0 (-5.8)* | 47.4 (5.4)** | 52.9 (32.6)** |
| Geometric mean travel times in minutes: | | | | | |
| Pharmacy, cycle | 9.4 (1.8)** | 7.7 (0.7)** | 6.7 (0.3)** | 6.3 (0.2)* | 5.7 (-0.4)** |
| Food stores, walk | 9.1 (1.8)** | 6.9 (0.9)** | 5.9 (0.6)** | 5.5 (0.5)** | 5.6 (-0.1) |
| Hospital, walk | 37.3 (6.5)** | 34.7 (2.4)** | 30.0 (1.6)** | 27.7 (0.5) | 25.7 (-2.0)** |
| GP, walk | 11.9 (1.5)** | 9.4 (0.7)** | 8.3 (0.2) | 7.7 (-0.2)* | 6.9 (-1.1)** |
| Further Education, walk | 20.1 (2.7)* | 17.9 (0.5)* | 16.0 (-0.2) | 15.5 (-0.6)* | 15.0 (-2.9)** |
| Large employment centres, walk | 36.3 (0.03) | 33.1 (3.1)** | 28.7 (-4.5)** | 24.3 (-2.9)** | 17.9 (-0.8) |
| Job Centre, public transport | 66.9 (0.1) | 63.6 (4.5)** | 59.2 (-7.6)** | 53.6 (-8.5)** | 47.9 (-12.8)** |
| Geometric mean distance to: | | | | | |
| Gambling shops | 1.8 (0.8)** | 1.2 (0.3)** | 0.9 (-0.1)** | 0.8 (-0.1)** | 0.6(0.02)** |
| Fast food shops | 1.9 (0.6)* | 1.2 (0.3)** | 0.9 (-0.1)* | 0.8 (0.00) | 0.6 (0.1)** |
| Pubs | 1.3 (0.3)** | 1.0 (0.3)** | 0.8 (-0.01) | 0.8 (0.03) | 0.7 (-0.1)** |
| Leisure | 3.2 (0.7)* | 2.5 (0.1) | 2.0 (-0.2)** | 1.8 (-0.3)** | 1.4 (-0.4)** |
| Tobacco shops | 4.2 (1.0)** | 2.9 (0.6)** | 2.0 (0.4)** | 1.7 (0.4)** | 1.5 (0.2) |
| Green space within 900m | 0.7 (0.1)** | 0.7 (0.2)** | 0.6 (0.2)** | 0.6 (0.1)** | 0.5 (0.1)** |
| Mean annual nitrogen dioxide (µgm$^3$) | 10.1 (-0.1)* | 10.6 (0.4)** | 11.3 (1.2)** | 12.1 (2.4)** | 13.1 (4.9)** |
| Mean annual Particulate Matter 10 (µgm$^3$) | 12.6 (0.3)** | 12.7 (0.5) | 12.8 (0.7)** | 12.7 (1.2)** | 12.6 (2.4)** |
| Mean annual Sulphur Dioxide (µgm$^3$) | 1.3 (-0.1)** | 1.3 (-0.1)** | 1.4 (-0.1)** | 1.4 (-0.1)** | 1.6 (-0.3)** |
| Social Environment: | | | | | |
| Mean % qualifications level 3+ | 40.6 (4.5)** | 36.8 (3.0)** | 33.5 (3.6)** | 29.0 (5.8)** | 25.2 (11.7)** |
| Mean % adults low skilled occupations | 12.3 (-1.1)** | 16.1 (-0.9)** | 19.8 (-1.8)** | 23.7 (-3.9)** | 26.2 (-8.2)** |
| Index of deprivation Crime quintile, 2015 | | | | | |
| Quintile 1 (ref – least deprived) | 41.5 (4.5) | 15.3 (1.8) | 4.2 (-0.5) | 1.0 (-0.1) | 0.0 (0.5) |
| Quintile 2 | 30.7 (-0.1) | 25.7 (2.4) | 13.2 (1.0) | 7.1 (-1.9) | 1.9 (0.3) |
| Quintile 3 | 18.4 (-2.3) | 28.5 (0.2) | 25.7 (3.1) | 17.1 (-1.9) | 7.1 (0.3) |
| Quintile 4 | 7.2 (-1.1) | 20.9 (-1.5) | 31.9 (0.8) | 25.0 (10.0) | 16.3 (9.6) |
| Quintile 5 (most deprived) | 2.2 (-1.0)** | 9.6 (-2.9)** | 25.0 (-4.4)** | 49.8 (-6.2)** | 74.6 (-10.6)** |
| Mean % age 10-19 years (SD) | 11.3 (0.7)** | 11.4 (0.3)** | 11.9 (-0.1) | 12.8 (-0.3)* | 13.0 (-0.2) |
| Economic Environment: | | | | | |
| Mean % unemployed 2011 (SD) | 2.5 (-0.2)** | 3.7 (-0.2)** | 5.1 (-0.3)** | 7.3 (-0.9)** | 10.3 (-2.3)** |
| Mean % overcrowded 2011 (SD) | 0.45 (0.06)* | 0.90 (0.22)* | 1.49 (0.59)** | 2.16 (1.92)** | 3.24 (5.01)** |
| Mean % no car 2011 (SD) | 12.2 (-2.4)** | 21.0 (-2.5)** | 31.2 (2.8)** | 43.4 (-4.0)** | 58.9 (-2.9)** |
| Mean % non-homeowner 2011 (SD) | 14.4 (2.3)** | 27.8 (1.7)** | 42.4 (-0.6)* | 58.5 (-3.9)** | 75.5 (-3.6)** |
| Mean % economically inactive 2011 (SD) | 31.3 (-3.1)** | 30.2 (-2.8)** | 31.3 (-2.6)** | 34.8 (-2.9)** | 40.0 (-5.0)** |
| Educational Environment: [a] | | | | | |
| Mean % 5+ A*-C grades, English & maths [b] | 63.4 (4.0)** | 56.8 (5.1)** | 51.7 (4.6)** | 46.6 (5.3)** | 40.9 (11.1)** |
| Mean total point score for MSOA (SD) [c] | 487 (3.8) | 472 (8.1)** | 461 (-10.0)** | 449 (10.2)** | 433 (16.2)** |
| POLAR higher education quintiles: | | | | | |
| Quintile 1 (ref – least participation) | 9.9 (-6.6) | 26.6 (-14.5) | 40.1 (-15.5) | 58.8 (-26.9) | 65.6 (-48.3) |
| Quintile 2 | 22.3 (-10.0) | 28.4 (-4.1) | 30.7 (-5.4) | 24.1 (-4.0) | 25.2 (-13.4) |
| Quintile 3 | 24.0 (-4.1) | 21.5 (-6.4) | 14.9 (2.4) | 9.5 (4.6) | 6.0 (15.8) |
| Quintile 4 | 24.9 (1.9) | 13.3 (-5.7) | 7.9 (5.3) | 2.6 (12.2) | 1.5 (32.5) |
| Quintile 5 (most participation) | 19.0 (18.8)** | 10.2 (12.8)** | 6.5 (13.2)** | 5.1 (14.2)** | 1.7 (13.4)** |

**Supplementary Table 4: P-value for interaction of each environmental predictor and coastal residence in adolescence in associations with SF-12 MCS, UKHLS youth sample, 2009-2021 (n=4,961)**

| | p-values for age-adjusted model | p-values for fully adjusted model |
|---|---|---|
| Economic Environment: | | |
| Mean % unemployed 2011 (SD) | **0.002** | 0.053 |
| Mean % overcrowded 2011 (SD) | 0.080 | 0.055 |
| Mean % no car 2011 (SD) | **0.004** | **0.043** |
| Mean % non-homeowner 2011 (SD) | **0.003** | **0.024** |
| Mean % economically inactive 2011 (SD) | 0.079 | 0.262 |
| Social Environment: | | |
| Mean % qualifications level 3+ (SD) | 0.342 | 0.288 |
| Mean % adults low skilled occupations (SD) | 0.565 | 0.449 |
| Index of deprivation Crime quintile, 2015 | | |
| Quintile 1 (ref – least deprived) | - | - |
| Quintile 2 | 0.711 | 0.838 |
| Quintile 3 | **0.049** | 0.075 |
| Quintile 4 | 0.317 | 0.866 |
| Quintile 5 (most deprived) | 0.076 | 0.413 |
| Mean % age 10-19 years (SD) | **0.001** | **0.007** |
| Educational Environment: [a] | | |
| % 5+ A*-C grades, English & maths | 0.226 | 0.190 |
| Mean capped point score for MSOA (SD) | **0.031** | **0.043** |
| POLAR participation rates | 0.454 | 0.206 |
| POLAR progress higher education quintiles: | | |
| Quintile 1 (ref – least progression) | - | - |
| Quintile 2 | 0.519 | 0.733 |
| Quintile 3 | 0.535 | 0.122 |
| Quintile 4 | 0.451 | 0.146 |
| Quintile 5 (most progression) | 0.810 | 0.799 |
| Built Environment: | | |
| Urban, % | 0.598 | 0.960 |
| Geometric mean persons per hectare (SD) | 0.073 | 0.433 |
| Geometric mean travel times in minutes (SD): | | |
| Pharmacy, cycle | **0.023** | 0.163 |
| Food stores, walk | 0.240 | 0.676 |
| Hospital, walk | 0.343 | 0.957 |
| GP, walk | **<0.001** | **<0.001** |
| Further Education, walk | **0.015** | 0.118 |
| Large employment centres, walk | **0.015** | 0.204 |
| Nearest job centre, public transport | **0.001** | **0.027** |
| AHAH measures (SD) | | |
| Geometric mean minutes to gambling shops | 0.270 | 0.785 |
| Geometric mean minutes to fast food shops | 0.166 | 0.642 |
| Geometric mean minutes to pubs | 0.262 | 0.790 |
| Geometric mean minutes to leisure | 0.059 | 0.296 |
| Geometric mean green space within 900m | 0.215 | 0.384 |
| Geometric mean minutes to tobacco shops | 0.053 | 0.283 |
| Mean annual nitrogen dioxide (µgm³) | **0.017** | 0.057 |
| Mean annual Particulate Matter 10 (µgm³) | 0.064 | 0.199 |
| Mean annual Sulphur Dioxide (µgm³) | 0.136 | 0.278 |

**Supplementary Table 3: P-value for interaction of each environmental predictor and coastal residence in adolescence in associations with SF-12 MCS, UKHLS youth sample, 2009-2021 (n=4,961)**

| | p-values for age-adjusted model | p-values for fully adjusted model |
|---|---|---|
| Economic Environment: | | |
| Mean % unemployed 2011 (SD) | **0.002** | 0.053 |
| Mean % overcrowded 2011 (SD) | 0.080 | 0.055 |
| Mean % no car 2011 (SD) | **0.004** | **0.043** |
| Mean % non-homeowner 2011 (SD) | **0.003** | **0.024** |
| Mean % economically inactive 2011 (SD) | 0.079 | 0.262 |
| Social Environment: | | |
| Mean % qualifications level 3+ (SD) | 0.342 | 0.288 |
| Mean % adults low skilled occupations (SD) | 0.565 | 0.449 |
| Index of deprivation Crime quintile, 2015 | | |
| Quintile 1 (ref – least deprived) | - | - |
| Quintile 2 | 0.711 | 0.838 |
| Quintile 3 | **0.049** | 0.075 |
| Quintile 4 | 0.317 | 0.866 |
| Quintile 5 (most deprived) | 0.076 | 0.413 |
| Mean % age 10-19 years (SD) | **0.001** | **0.007** |
| Educational Environment: [a] | | |
| % 5+ A*-C grades, English & maths | 0.226 | 0.190 |
| Mean capped point score for MSOA (SD) | **0.031** | **0.043** |
| POLAR participation rates | 0.454 | 0.206 |
| POLAR progress higher education quintiles: | | |
| Quintile 1 (ref – least progression) | - | - |
| Quintile 2 | 0.519 | 0.733 |
| Quintile 3 | 0.535 | 0.122 |
| Quintile 4 | 0.451 | 0.146 |
| Quintile 5 (most progression) | 0.810 | 0.799 |
| Built Environment: | | |
| Urban, % | 0.598 | 0.960 |
| Geometric mean persons per hectare (SD) | 0.073 | 0.433 |
| Geometric mean travel times in minutes (SD): | | |
| Pharmacy, cycle | **0.023** | 0.163 |
| Food stores, walk | 0.240 | 0.676 |
| Hospital, walk | 0.343 | 0.957 |
| GP, walk | **<0.001** | **<0.001** |
| Further Education, walk | **0.015** | 0.118 |
| Large employment centres, walk | **0.015** | 0.204 |
| Nearest job centre, public transport | **0.001** | **0.027** |
| AHAH measures (SD) | | |
| Geometric mean minutes to gambling shops | 0.270 | 0.785 |
| Geometric mean minutes to fast food shops | 0.166 | 0.642 |
| Geometric mean minutes to pubs | 0.262 | 0.790 |
| Geometric mean minutes to leisure | 0.059 | 0.296 |
| Geometric mean green space within 900m | 0.215 | 0.384 |
| Geometric mean minutes to tobacco shops | 0.053 | 0.283 |
| Mean annual nitrogen dioxide ($\mu gm^3$) | **0.017** | 0.057 |
| Mean annual Particulate Matter 10 ($\mu gm^3$) | 0.064 | 0.199 |
| Mean annual Sulphur Dioxide ($\mu gm^3$) | 0.136 | 0.278 |
